# Contribution of cerebrovascular lesions to neurodegeneration and cognitive decline

**DOI:** 10.64898/2026.10.04.26364698

**Authors:** Zhaojin Chen, Shima Raeesi, Amelie Metz, Roqaie Moqadam, Katherine Chadwick, the Consortium for the early identification of Alzheimer’s disease–Quebec (CIMA-Q), Yashar Zeighami, Mahsa Dadar

**Affiliations:** Douglas Research Center, Montréal, Québec, Canada; Integrated Program in Neuroscience, McGill University, Montréal, Québec, Canada; Department of Medicine, Université de Montréal, Montréal, Québec, Canada; Department of Psychiatry, McGill University, Montréal, Québec, Canada

**Author notes:** Corresponding authors: Zhaojin Chen,; Mahsa Dadar,; Yashar Zeighami, Address: Douglas Mental Health University Institute, 6875 Bd LaSalle CIC Building, Verdun, QC H4H 1R3. Data used in the preparation of this article were obtained from the Consortium for the early identification of Alzheimer’s disease – Quebec (CIMA-Q; cima-q.ca). A list of researchers involved in the design of CIMA-Q can be found on the cima-q.ca website. These researchers contributed to the establishment of protocols, the implementation of the research infrastructure, the recruitment and follow-up of participants, the obtaining of data, the maintenance of biological and ex-vivo samples, and certain derived data.

**Keywords:** Cerebral small vessel disease, magnetic resonance imaging, white matter hyperintensities, microbleeds, lacunar infarcts, perivascular spaces, cognition, Alzheimer’s disease

## Abstract

**Introduction:** Cerebral small vessel disease (CSVD) lesions, including white matter hyperintensities (WMHs), lacunar infarcts, cerebral microbleeds (CMBs), and enlarged perivascular spaces (PVSs) are associated with increased risk of dementia. In this study, we aimed to examine their associations with risk factors, neurodegeneration, and cognition over a follow-up period of ten years.

**Methods:** 289 participants (183 females, mean age: 73.7 ± 5.7) with multi-modal magnetic resonance imaging (MRI) and clinical assessments from the Consortium for the Early Identification of Alzheimer’s Disease (CIMA-Q) were included. CSVD lesions and neurodegeneration were quantified using automated tools. Linear regression and mixed effects models examined the cross-sectional and longitudinal associations between lesions, risk factors, neurodegeneration, and cognitive decline.

**Results:** Risk factors showed the strongest cross-sectional associations with PVS number and CMB presence, and the strongest longitudinal associations with WMH and PVS volume progression *(p < 0.05)*. WMH and PVS volumes significantly increased over time (p < 0.01), with faster WMH progression among participants who experienced cognitive decline during study follow-ups. Higher WMH volume was associated with poorer global cognition and executive function, and presence of lacunae was associated with faster decline in executive function. Finally, both WMH and PVS were associated with longitudinal atrophy across several cortical regions.

**Conclusions:** Vascular risk factors were associated with CSVD burden and progression, and WMH and PVS progression were further associated with cortical atrophy and cognitive decline, underscoring the interconnected contributions of vascular risk, CSVD, and neurodegeneration to cognitive aging and dementia risk.

**HIGHLIGHT:**

- White matter hyperintensities (WMHs) and perivascular spaces (PVS) showed significant longitudinal increases over up to 10 years of study follow-ups.
- Vascular risk factors were associated with CSVD burden (PVSs, cerebral microbleeds, and WMHs) and progression (PVSs and WMHs).
- Greater WMH burden was associated with poorer global cognition and executive function, while lacunar infarcts were associated with faster longitudinal decline in executive function.
- WMH and PVS progression were associated with longitudinal cortical atrophy, linking CSVD progression with neurodegenerative changes.

## 1 BACKGROUND

Cerebral small vessel disease (CSVD) is the most common vascular contributor to dementia, and a frequent co-pathology in individuals on the Alzheimer’s disease (AD) continuum^1–3^. Many of the risk factors that are associated with AD are also known contributors to CSVD, with even more direct links to vascular processes (e.g. hypertension, smoking, and excessive alcohol consumption)^4–6^. CSVD can be heterogeneous in different populations, with varying presentations based on their specific genetic predisposition and risk factor profiles^7–10^. Accumulating evidence suggests that AD and vascular pathologies can interact and exacerbate each other, leading to worsening cognitive decline and clinical outcomes^11–14^.

The primary markers of CSVD can be identified using magnetic resonance imaging (MRI), including white matter hyperintensities (WMH), cerebral microbleeds, lacunae, and enlarged perivascular spaces (PVSs)^1^, which reflect partially distinct vascular and pathological processes. WMHs are defined as areas of hyperintense signal on T2-weighted (T2w) or fluid attenuated inversion recovery (FLAIR) MRIs^1,15^. The iron signal corresponding to cerebral microbleeds is detectable on T2* or susceptibility weighted images (SWI)^1,16^, and lacunae and PVSs can be detected using multi-modal combinations of T1-weighted (T1w) and T2w or FLAIR MRIs^1^.

While WMHs have a presumed vascular origin, recent literature suggests that WMHs, in particular in posterior brain regions, can be more strongly linked to downstream AD pathology than vascular processes in individuals with AD^17–20^. Regardless, WMHs are very commonly observed in aging individuals (with prevalence rates for moderate-to-severe WMHs ranging from 13-48%)^9,21,22^ and have been linked to future neurodegenerative changes, cortical thinning, cognitive impairment and dementia, as well as neuropsychiatric symptoms (e.g. depression)^13,23–26^. Lacunae have a more direct vascular origin, with reported prevalence rates ranging from 8-28% in general aging populations, and have also been linked to cognitive impairment and dementia^22,27–29^. Microbleeds can be asymptomatic or linked to cognitive deficits and dementia in normal aging and AD, and can result from both vascular processes and amyloid deposition, particularly in individuals with AD^2,30,31^. Finally, enlarged MRI-visible PVSs have been associated with worsening cognitive function and increased risk of dementia in some populations^2,32,33^.

In the past decade, FLAIR imaging has been consistently added to MRI protocols in most aging and AD datasets^34–37^. With the recent advances in reliable fully automated WMH quantification methods^38^, numerous studies have examined the role of WMHs in AD, and their association with amyloid and tau pathologies, neurodegeneration, and cognitive decline^11–13,39,40^. In contrast, T2* and SWI sequences are less frequently and consistently available, and current automated microbleed and lacune segmentation methods still require extensive additional quality control and manual correction, to ensure accuracy of the derived measures^38^. As such, despite the clinical relevance of CSVD lesions, few studies have simultaneously characterized their prevalence, longitudinal progression trajectories, and associations with neurodegeneration and cognitive decline in cohorts with large sample sizes and long-term longitudinal follow-ups^41–44^. Consequently, most longitudinal studies have focused on individual CSVD markers, particularly WMHs, rather than simultaneously characterizing multiple CSVD manifestations. This limits our understanding of whether different CSVD lesions follow distinct trajectories and make independent or complementary contributions to neurodegeneration and cognitive decline.

Taking advantage of the longitudinal multi-modal MRI protocol of the Consortium for the Early Identification of Alzheimer’s Disease (CIMA-Q), allowing for longitudinal quantification of all four CSVD lesion types in 289 individuals followed for up to 10 years, the present study investigates the prevalence and longitudinal trajectories of CSVD lesions in individuals on the AD continuum. We further examine their associations with neurodegeneration and cognitive decline to determine the extent to which distinct manifestations of CSVD contribute to disease progression.

## 2 METHODS

### 2.1 Participants

Data were obtained from the CIMA-Q^35^, a multicenter, longitudinal cohort study. Most participants were recruited in Quebec through advertisements and memory clinics. Participants underwent standardized neuropsychological and neuropsychiatric evaluations conducted by CIMA-Q–certified psychometricians and completed questionnaires covering health, lifestyle, and other relevant factors. Clinical diagnoses were established by expert physicians according to the National Institute on Aging–Alzheimer’s Association (NIA-AA) diagnostic criteria^45^. Only participants who underwent MRI acquisition were included in this study. Three participants were additionally excluded as their final diagnosis fell outside the diagnostic categories of interest.

### 2.2 Consensus Diagnosis

Consensus diagnoses were defined by a geriatrician (SR) based on all available longitudinal diagnosis information, as well as cognitive and functional status assessed by the MoCA and CDR sum of boxes (CDR-SB), and available domain-specific cognitive measures, including verbal memory and language performance. In brief, participants were categorized as cognitively intact controls if they had a consistent clinical diagnosis of cognitively normal/control (CN) across longitudinal visits. Similarly, participants with subjective cognitive decline (SCD), mild cognitive impairment (MCI) and AD were classified as stable SCD, stable MCI, and stable AD, respectively, if their diagnoses remained consistent across longitudinal visits. Participants were classified as converter-SCD and converter-MCI if they were subsequently diagnosed with MCI (e.g. SCD → SCD → MCI → MCI → MCI) or dementia (e.g. MCI → MCI → MCI → AD →AD) during follow-ups, respectively. Inconsistent diagnoses were re-assigned based on the most stable diagnoses and cognitive scores. For example, SCD → SCD → MCI → SCD → SCD was classified as stable SCD. To increase statistical power, converter-SCD and converter-MCI participants were combined into a converter group as both groups experienced progression to a more impaired diagnostic category during follow-ups.

### 2.3 Image Processing

PELICAN pipeline^46^ was used to process T1w, FLAIR, and T2w images. Preprocessing steps included denoising, bias-field correction, and intensity normalization. FLAIR and T2w images were then co-registered to the corresponding T1w images, followed by longitudinal linear and nonlinear registration to the MNI-ICBM-2009c average template^47,48^. T2*-weighted images were co-registered to the corresponding T2w images using ANTs^48^ and subsequently transformed to the MNI-ICBM-2009c space. Tissue segmentation (including WMHs) was automatically performed based on T1w and FLAIR images using the BISON tool^49,50^. Voxel-based morphometry (VBM) maps were generated by modulating the Jacobian determinant of the nonlinear deformation fields based on BISON tissue probability maps^51,52^. Regional gray matter (GM) VBM measures were subsequently extracted using the CerebrA atlas^47^. CSVD lesions, including cerebral microbleeds (CMBs), lacunae, and PVSs were automatically segmented using ShiVA tools^53–55^ based on T2*-weighted, FLAIR, and T1w images, respectively. All registrations and segmentations were visually quality controlled by expert raters. Segmentation errors for WMHs, microbleeds, and lacunae were manually corrected. For PVSs, cases that failed segmentation were excluded from subsequent analyses. Cases that failed nonlinear registration were excluded from the VBM analyses. The final segmentation maps were transformed into the MNI-ICBM-2009c space. Figure 1 shows an example of the derived segmentations for a CIMA-Q participant. WMH, PVS, CMB, and lacunae volumes were quantified by summing the voxels within their respective segmentation masks and are reported in cubic millimeters (mm³). PVS, CMB, and lacunae counts were also extracted.

**Figure 1.**
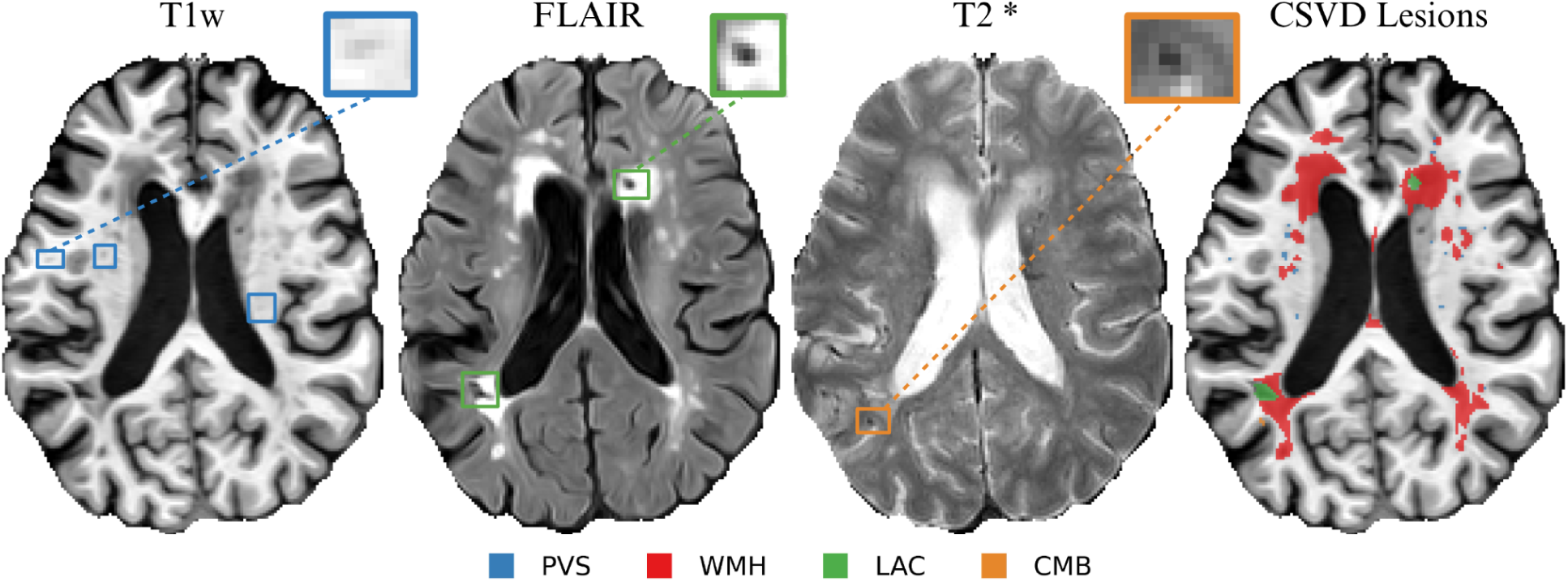
CSVD lesion segmentations in a CIMA-Q participant. CSVD: Cerebral small vessel disease. PVS: Perivascular spaces. CMB: Cerebral microbleed. WMH: White Matter Hyperintensity. LAC: Lacunae.

### 2.4 Vascular Health Indicators

To comprehensively evaluate the association of vascular risk with cerebrovascular lesions, we assessed clinical diagnoses, lifestyle factors, anthropometric measures, blood pressure, and laboratory biomarkers. These indicators were selected to capture complementary aspects of vascular health, including established cardiometabolic conditions, glucose and lipid metabolism, and adiposity distribution. Hypertension, diabetes, and hypercholesterolemia were defined using the corresponding clinical diagnosis variables. Smoking history was determined by self-reported lifetime smoking of at least 100 cigarettes, whereas alcohol consumption was assessed based on self-reported current alcohol use. Current smoking status was additionally used to calculate the Framingham risk score. Physical inactivity was defined as reporting no current physical activity. These categorical indicators were coded as absent (0) or present (1).

Continuous anthropometric indicators included body mass index (BMI), calculated as weight in kilograms divided by height in meters squared (kg/m²); neck, waist, and hip circumferences; and waist-to-hip ratio, calculated by dividing waist circumference by hip circumference. These measures were examined separately because BMI reflects overall adiposity but does not characterize regional fat distribution. Waist circumference and waist-to-hip ratio were included as indicators of central adiposity; hip circumference was retained to characterize body-fat distribution and calculate the waist-to-hip ratio; and neck circumference was examined as a complementary indicator of upper-body adiposity and associated cardiometabolic and sleep-apnea risk. Additional continuous indicators included resting systolic and diastolic blood pressure, glycated hemoglobin (HbA1c), total cholesterol, triglycerides, low-density lipoprotein (LDL) cholesterol, and high-density lipoprotein (HDL) cholesterol.

### 2.5 Vascular Composite Scores

An unweighted vascular composite score was used to characterize cumulative vascular risk burden^56,57^. The score included six clinically relevant risk factors: hypertension, diabetes, hypercholesterolemia, smoking, high BMI (≥30 kg/m²), and physical inactivity. Each component was coded as present (1) or absent (0), and the components were summed to produce a score ranging from 0 to 6, with higher scores indicating greater cumulative vascular risk burden. The score was calculated only for participants with complete data for all six components. This additive approach provides a transparent and clinically interpretable measure of cumulative vascular risk, avoids data-driven weighting and potential overfitting, and preserves the contribution of each individual risk factor.

As a complementary weighted measure, we calculated the lipid-based Framingham continuous model for 10-year general cardiovascular disease risk^58,59^. The model incorporates age, sex, total cholesterol, HDL cholesterol, systolic blood pressure, antihypertensive treatment, diabetes, and current-smoking status. Because antihypertensive medication data were unavailable, participants with a diagnosis of hypertension were assumed to be receiving antihypertensive treatment, whereas those without hypertension were classified as untreated. The sex-specific continuous-model linear predictor was calculated using the published Framingham coefficients, with higher values indicating greater estimated cardiovascular risk. The score was calculated only for participants with complete data for all required components.

The Hachinski Ischemic Score was additionally examined as an established clinical index of vascular involvement. Unlike the preceding vascular-risk measures, it evaluates clinical features suggestive of a cerebrovascular contribution to cognitive impairment, with higher scores indicating stronger evidence of vascular involvement. Raw values for all three scores are presented in Table 1, and each score was standardized before inclusion in the regression models.

**Table 1.**
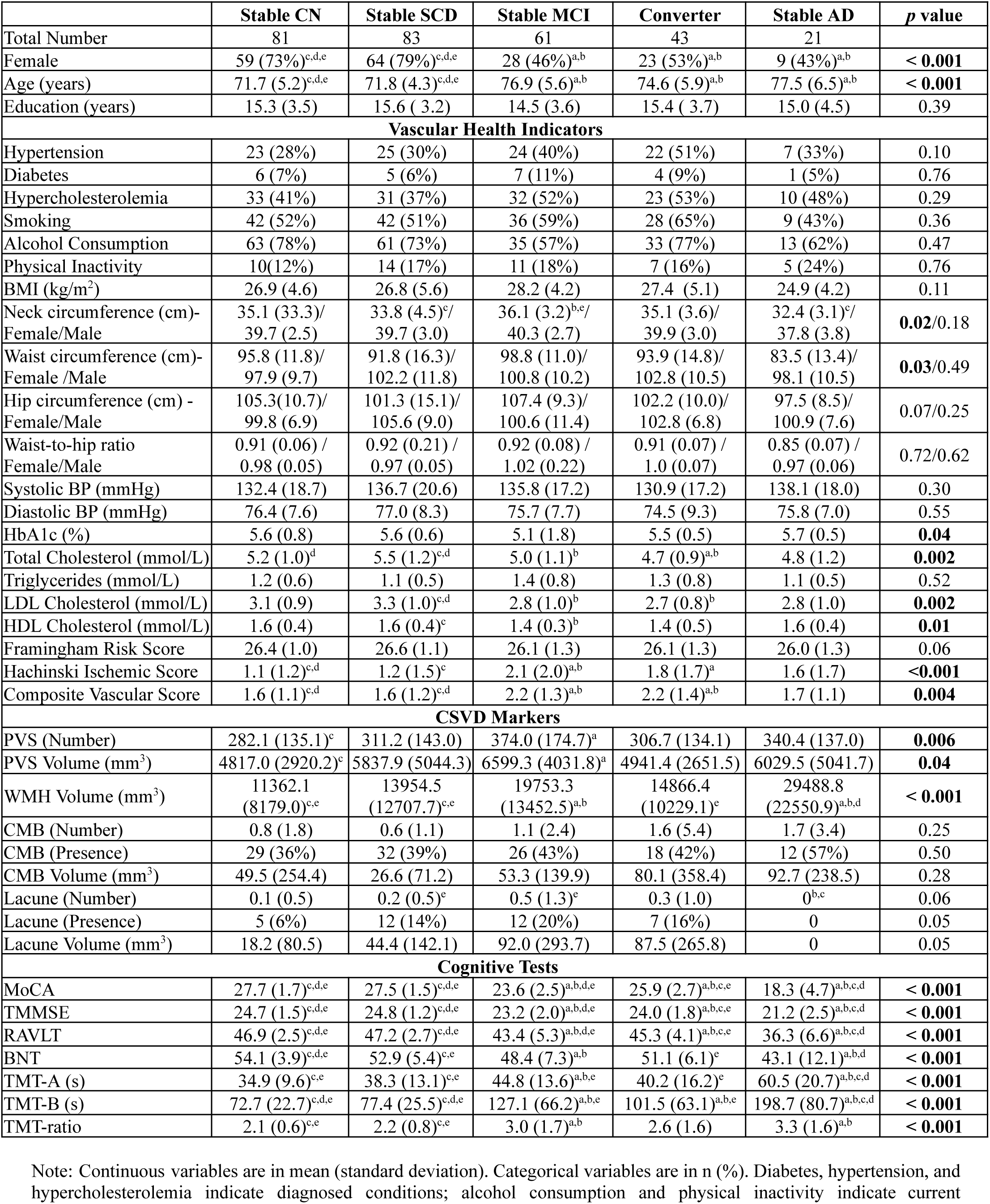
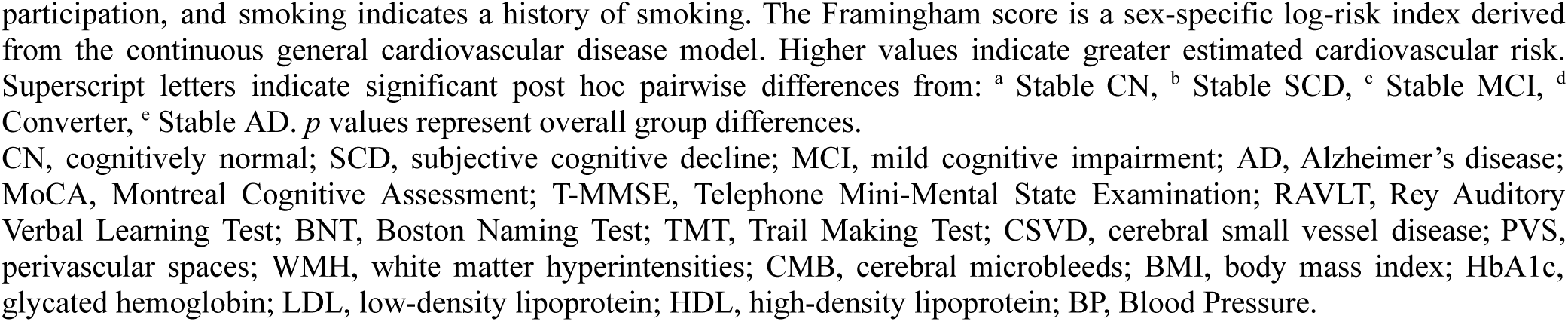
Participant characteristics at the first available assessment by diagnostic group.

|  | Stable CN | Stable SCD | Stable MCI | Converter | Stable AD | *p* value |
| --- | --- | --- | --- | --- | --- | --- |
| Total Number | 81 | 83 | 61 | 43 | 21 |  |
| Female | 59 (73%)<sup>c,d,e</sup> | 64 (79%)<sup>c,d,e</sup> | 28 (46%)<sup>a,b</sup> | 23 (53%)<sup>a,b</sup> | 9 (43%)<sup>a,b</sup> | **< 0.001** |
| Age (years) | 71.7 (5.2)<sup>c,d,e</sup> | 71.8 (4.3)<sup>c,d,e</sup> | 76.9 (5.6)<sup>a,b</sup> | 74.6 (5.9)<sup>a,b</sup> | 77.5 (6.5)<sup>a,b</sup> | **< 0.001** |
| Education (years) | 15.3 (3.5) | 15.6 (3.2) | 14.5 (3.6) | 15.4 (3.7) | 15.0 (4.5) | 0.39 |
| **Vascular Health Indicators** |  |  |  |  |  |  |
| Hypertension | 23 (28%) | 25 (30%) | 24 (40%) | 22 (51%) | 7 (33%) | 0.10 |
| Diabetes | 6 (7%) | 5 (6%) | 7 (11%) | 4 (9%) | 1 (5%) | 0.76 |
| Hypercholesterolemia | 33 (41%) | 31 (37%) | 32 (52%) | 23 (53%) | 10 (48%) | 0.29 |
| Smoking | 42 (52%) | 42 (51%) | 36 (59%) | 28 (65%) | 9 (43%) | 0.36 |
| Alcohol Consumption | 63 (78%) | 61 (73%) | 35 (57%) | 33 (77%) | 13 (62%) | 0.47 |
| Physical Inactivity | 10(12%) | 14 (17%) | 11 (18%) | 7 (16%) | 5 (24%) | 0.76 |
| BMI (kg/m<sup>2</sup>) | 26.9 (4.6) | 26.8 (5.6) | 28.2 (4.2) | 27.4 (5.1) | 24.9 (4.2) | 0.11 |
| Neck circumference (cm)-<br>Female/Male | 35.1 (33.3)/<br>39.7 (2.5) | 33.8 (4.5)<sup>c</sup>/<br>39.7 (3.0) | 36.1 (3.2)<sup>b,e</sup>/<br>40.3 (2.7) | 35.1 (3.6)/<br>39.9 (3.0) | 32.4 (3.1)<sup>c</sup>/<br>37.8 (3.8) | **0.02/0.18** |
| Waist circumference (cm)-<br>Female /Male | 95.8 (11.8)/<br>97.9 (9.7) | 91.8 (16.3)/<br>102.2 (11.8) | 98.8 (11.0)/<br>100.8 (10.2) | 93.9 (14.8)/<br>102.8 (10.5) | 83.5 (13.4)/<br>98.1 (10.5) | **0.03/0.49** |
| Hip circumference (cm) -<br>Female/Male | 105.3(10.7)/<br>99.8 (6.9) | 101.3 (15.1)/<br>105.6 (9.0) | 107.4 (9.3)/<br>100.6 (11.4) | 102.2 (10.0)/<br>102.8 (6.8) | 97.5 (8.5)/<br>100.9 (7.6) | 0.07/0.25 |
| Waist-to-hip ratio<br>Female/Male | 0.91 (0.06) /<br>0.98 (0.05) | 0.92 (0.21) /<br>0.97 (0.05) | 0.92 (0.08) /<br>1.02 (0.22) | 0.91 (0.07) /<br>1.0 (0.07) | 0.85 (0.07) /<br>0.97 (0.06) | 0.72/0.62 |
| Systolic BP (mmHg) | 132.4 (18.7) | 136.7 (20.6) | 135.8 (17.2) | 130.9 (17.2) | 138.1 (18.0) | 0.30 |
| Diastolic BP (mmHg) | 76.4 (7.6) | 77.0 (8.3) | 75.7 (7.7) | 74.5 (9.3) | 75.8 (7.0) | 0.55 |
| HbA1c (%) | 5.6 (0.8) | 5.6 (0.6) | 5.1 (1.8) | 5.5 (0.5) | 5.7 (0.5) | **0.04** |
| Total Cholesterol (mmol/L) | 5.2 (1.0)<sup>d</sup> | 5.5 (1.2)<sup>c,d</sup> | 5.0 (1.1)<sup>b</sup> | 4.7 (0.9)<sup>a,b</sup> | 4.8 (1.2) | **0.002** |
| Triglycerides (mmol/L) | 1.2 (0.6) | 1.1 (0.5) | 1.4 (0.8) | 1.3 (0.8) | 1.1 (0.5) | 0.52 |
| LDL Cholesterol (mmol/L) | 3.1 (0.9) | 3.3 (1.0)<sup>c,d</sup> | 2.8 (1.0)<sup>b</sup> | 2.7 (0.8)<sup>b</sup> | 2.8 (1.0) | **0.002** |
| HDL Cholesterol (mmol/L) | 1.6 (0.4) | 1.6 (0.4)<sup>c</sup> | 1.4 (0.3)<sup>b</sup> | 1.4 (0.5) | 1.6 (0.4) | **0.01** |
| Framingham Risk Score | 26.4 (1.0) | 26.6 (1.1) | 26.1 (1.3) | 26.1 (1.3) | 26.0 (1.3) | 0.06 |
| Hachinski Ischemic Score | 1.1 (1.2)<sup>c,d</sup> | 1.2 (1.5)<sup>c</sup> | 2.1 (2.0)<sup>a,b</sup> | 1.8 (1.7)<sup>a</sup> | 1.6 (1.7) | **<0.001** |
| Composite Vascular Score | 1.6 (1.1)<sup>c,d</sup> | 1.6 (1.2)<sup>c,d</sup> | 2.2 (1.3)<sup>a,b</sup> | 2.2 (1.4)<sup>a,b</sup> | 1.7 (1.1) | **0.004** |
| **CSVD Markers** |  |  |  |  |  |  |
| PVS (Number) | 282.1 (135.1)<sup>c</sup> | 311.2 (143.0) | 374.0 (174.7)<sup>a</sup> | 306.7 (134.1) | 340.4 (137.0) | **0.006** |
| PVS Volume (mm<sup>3</sup>) | 4817.0 (2920.2)<sup>c</sup> | 5837.9 (5044.3) | 6599.3 (4031.8)<sup>a</sup> | 4941.4 (2651.5) | 6029.5 (5041.7) | **0.04** |
| WMH Volume (mm<sup>3</sup>) | 11362.1<br>(8179.0)<sup>c,e</sup> | 13954.5<br>(12707.7)<sup>c,e</sup> | 19753.3<br>(13452.5)<sup>a,b</sup> | 14866.4<br>(10229.1)<sup>e</sup> | 29488.8<br>(22550.9)<sup>a,b,d</sup> | **< 0.001** |
| CMB (Number) | 0.8 (1.8) | 0.6 (1.1) | 1.1 (2.4) | 1.6 (5.4) | 1.7 (3.4) | 0.25 |
| CMB (Presence) | 29 (36%) | 32 (39%) | 26 (43%) | 18 (42%) | 12 (57%) | 0.50 |
| CMB Volume (mm<sup>3</sup>) | 49.5 (254.4) | 26.6 (71.2) | 53.3 (139.9) | 80.1 (358.4) | 92.7 (238.5) | 0.28 |
| Lacune (Number) | 0.1 (0.5) | 0.2 (0.5)<sup>e</sup> | 0.5 (1.3)<sup>e</sup> | 0.3 (1.0) | 0<sup>b,c</sup> | 0.06 |
| Lacune (Presence) | 5 (6%) | 12 (14%) | 12 (20%) | 7 (16%) | 0 | 0.05 |
| Lacune Volume (mm<sup>3</sup>) | 18.2 (80.5) | 44.4 (142.1) | 92.0 (293.7) | 87.5 (265.8) | 0 | 0.05 |
| **Cognitive Tests** |  |  |  |  |  |  |
| MoCA | 27.7 (1.7)<sup>c,d,e</sup> | 27.5 (1.5)<sup>c,d,e</sup> | 23.6 (2.5)<sup>a,b,d,e</sup> | 25.9 (2.7)<sup>a,b,c,e</sup> | 18.3 (4.7)<sup>a,b,c,d</sup> | **< 0.001** |
| TMMSE | 24.7 (1.5)<sup>c,d,e</sup> | 24.8 (1.2)<sup>c,d,e</sup> | 23.2 (2.0)<sup>a,b,d,e</sup> | 24.0 (1.8)<sup>a,b,c,e</sup> | 21.2 (2.5)<sup>a,b,c,d</sup> | **< 0.001** |
| RAVLT | 46.9 (2.5)<sup>c,d,e</sup> | 47.2 (2.7)<sup>c,d,e</sup> | 43.4 (5.3)<sup>a,b,d,e</sup> | 45.3 (4.1)<sup>a,b,c,e</sup> | 36.3 (6.6)<sup>a,b,c,d</sup> | **< 0.001** |
| BNT | 54.1 (3.9)<sup>c,d,e</sup> | 52.9 (5.4)<sup>c,e</sup> | 48.4 (7.3)<sup>a,b</sup> | 51.1 (6.1)<sup>e</sup> | 43.1 (12.1)<sup>a,b,d</sup> | **< 0.001** |
| TMT-A (s) | 34.9 (9.6)<sup>c,e</sup> | 38.3 (13.1)<sup>c,e</sup> | 44.8 (13.6)<sup>a,b,e</sup> | 40.2 (16.2)<sup>e</sup> | 60.5 (20.7)<sup>a,b,c,d</sup> | **< 0.001** |
| TMT-B (s) | 72.7 (22.7)<sup>c,d,e</sup> | 77.4 (25.5)<sup>c,d,e</sup> | 127.1 (66.2)<sup>a,b,e</sup> | 101.5 (63.1)<sup>a,b,e</sup> | 198.7 (80.7)<sup>a,b,c,d</sup> | **< 0.001** |
| TMT-ratio | 2.1 (0.6)<sup>c,e</sup> | 2.2 (0.8)<sup>c,e</sup> | 3.0 (1.7)<sup>a,b</sup> | 2.6 (1.6) | 3.3 (1.6)<sup>a,b</sup> | **< 0.001** |

Note: Continuous variables are in mean (standard deviation). Categorical variables are in n (%). Diabetes, hypertension, and hypercholesterolemia indicate diagnosed conditions; alcohol consumption and physical inactivity indicate current
participation, and smoking indicates a history of smoking. The Framingham score is a sex-specific log-risk index derived from the continuous general cardiovascular disease model. Higher values indicate greater estimated cardiovascular risk. Superscript letters indicate significant post hoc pairwise differences from: <sup>a</sup> Stable CN, <sup>b</sup> Stable SCD, <sup>c</sup> Stable MCI, <sup>d</sup> Converter, <sup>e</sup> Stable AD. *p* values represent overall group differences.
CN, cognitively normal; SCD, subjective cognitive decline; MCI, mild cognitive impairment; AD, Alzheimer's disease; MoCA, Montreal Cognitive Assessment; T-MMSE, Telephone Mini-Mental State Examination; RAVLT, Rey Auditory Verbal Learning Test; BNT, Boston Naming Test; TMT, Trail Making Test; CSVD, cerebral small vessel disease; PVS, perivascular spaces; WMH, white matter hyperintensities; CMB, cerebral microbleeds; BMI, body mass index; HbA1c, glycated hemoglobin; LDL, low-density lipoprotein; HDL, high-density lipoprotein; BP, Blood Pressure.

### 2.6 Cognitive Assessments

Participants completed a series of cognitive tests at baseline and scheduled follow-up visits. Assessments were administered in French or English using standardized instructions^35^. Cognitive outcomes included measures of global cognition, verbal episodic memory, processing speed, executive function, and language. Global cognition was assessed using the Montreal Cognitive Assessment (MoCA) and the 26-point Telephone Mini-Mental State Examination (T-MMSE). Verbal learning and episodic memory were assessed using the Rey Auditory Verbal Learning Test (RAVLT), in which participants learned a 15-word list over repeated trials and recalled the words after a delay. Processing speed and executive function were evaluated using the Trail Making Test (TMT). Part A primarily assesses processing speed, whereas Part B additionally assesses cognitive flexibility and set-shifting. The TMT B/A ratio was calculated by dividing the Part B completion time by the Part A completion time, providing an index of executive control while reducing the influence of processing speed. Language was assessed using the Boston Naming Test (BNT), a measure of visual confrontation naming. For all cognitive tests, higher scores indicated better performance. Because longer completion times and a higher B/A ratio on the TMT indicate poorer performance, TMT measures were reverse-coded in all analyses, except in Table 1, to ensure that higher scores consistently reflected better performance.

### 2.7 Statistical Analysis

Cross-sectional analyses used the first available observation for each participant, prioritizing the baseline visit and using the next available visit when baseline data were missing. Continuous variables are summarized as means and standard deviations, and categorical variables as frequencies and percentages. Differences across diagnostic groups were assessed using one-way analysis of variance (ANOVA) for continuous variables and Pearson’s chi-square or Fisher’s exact tests, as appropriate, for categorical variables. Overall significance tests in the demographics table were followed by pairwise group comparisons and false discovery rate (FDR) correction for multiple comparisons. Because body circumferences differ systematically by sex, comparisons of neck, waist, and hip circumferences were performed separately by sex. CSVD outcomes of interest included WMH volume; PVS count and volume; CMB count, volume, and presence; and lacune count, volume, and presence. CMB and lacune presence were coded as the presence of at least one lesion. Right-skewed lesion volumes and counts were log-transformed where appropriate, and continuous variables were standardized (z-scored) before inclusion in regression models. Values in Table 1 are presented in their original units.

Linear regression models examined the cross-sectional associations between variables of interest, whereas mixed-effects models examined the rate of progression in CSVD lesions as well as their longitudinal associations with neurodegeneration and cognition. For all longitudinal analyses, the Stable AD group was excluded because of insufficient repeated observations available.

Associations between vascular health indicators and CSVD lesion measures were first evaluated cross-sectionally using separate covariate-adjusted models for each vascular indicator and lesion outcome:

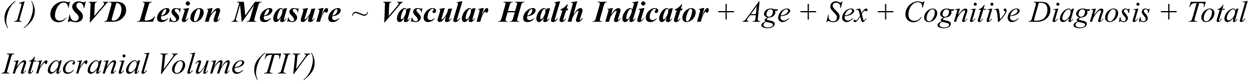

Where the variable of interest was *Vascular Health Indicator* (indicated in bold font), indicating lifestyle and anthropometric measures, blood-pressure and laboratory measures, the vascular composite score, the Framingham risk score, and the Hachinski Ischemic Score. Linear regressions were used for continuous lesion volumes (WMH and PVS), whereas logistic regressions were used for CMB and lacune presence. Continuous lesion volumes (WMH and PVS) were used as outcome variables and continuous vascular indicators as predictors in the following mixed-effects models to examine their longitudinal associations:

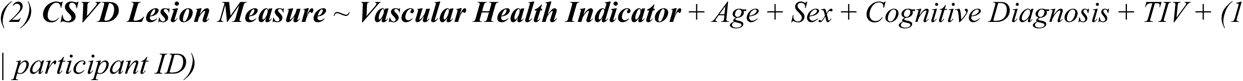

Longitudinal associations with binary vascular indicators were also evaluated using mixed-effects models, including the following vascular indicator-by-time interactions:

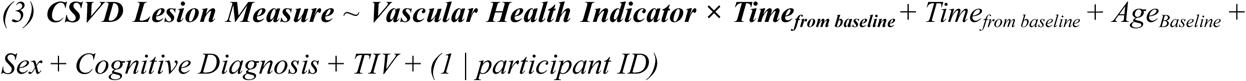

The interaction term tested whether lesion trajectories differed according to the presence of the vascular indicator. Associations among CSVD markers were examined using pairwise models in which each lesion marker was treated in turn as the outcome and the other marker as the predictor. Cross-sectional linear regression models were used for continuous lesion measures, and logistic regression models were used when CMB or lacune presence was the outcome of interest:

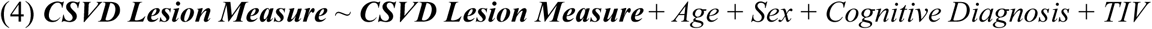

Longitudinal associations between WMH and PVS volumes across repeated assessments were examined using linear mixed-effects models with participant-specific random intercepts:

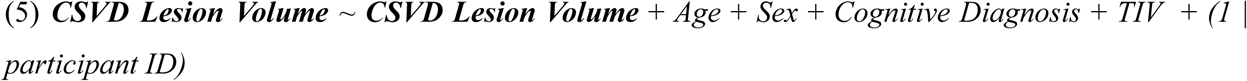

Separate interaction models tested whether baseline CMB or lacune presence was associated with subsequent rates of change in WMH or PVS volume:

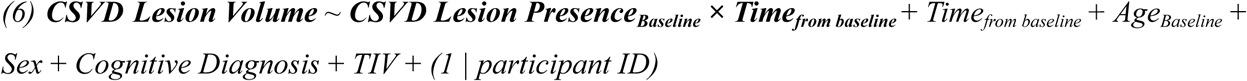

Longitudinal trajectories of WMH and PVS volumes and potential progression differences between diagnostic groups were examined using the following linear mixed-effects models:

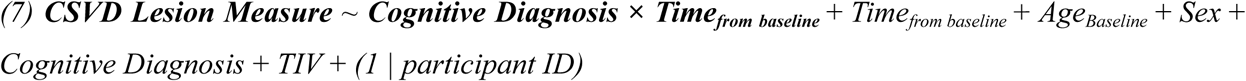

The time-by-diagnostic-group interaction tested whether rates of lesion volume change differed across groups. Associations between CSVD lesions and cognition were evaluated separately for each cognitive outcome. Cross-sectional linear regression models included WMH or PVS volume or CMB or lacune presence as the predictor and were adjusted for age, sex, cognitive diagnosis, TIV, and education:

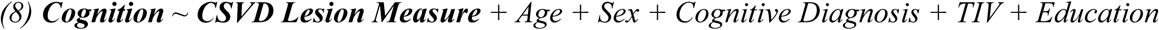

Longitudinal associations of time-varying WMH and PVS volumes with cognitive performance across repeated assessments were examined using linear mixed-effects models with the same covariates and a participant-specific random intercept.

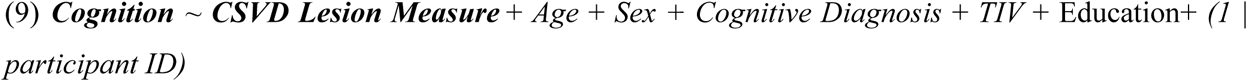

For CMBs and lacunae, interaction models were used to test whether lesion presence was associated with differences in longitudinal cognitive trajectories.

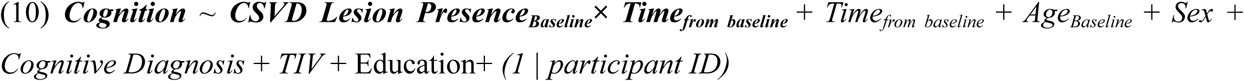

Associations between CSVD lesions and regional GM volumes were examined using VBM-derived regional volumes as outcomes. Cross-sectional linear models examined WMH volume, PVS volume, PVS count, CMB presence, and lacune presence separately:

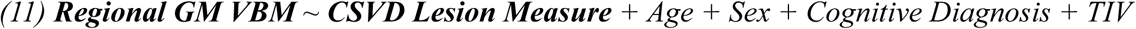

Longitudinal mixed-effects models examined the associations of WMH and PVS volumes with regional VBM volume across repeated assessments:

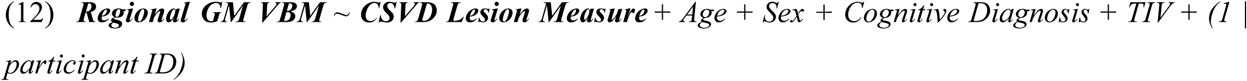

Additional models included baseline CMB- or lacune-presence-by-time interactions to evaluate their associations with subsequent regional GM VBM trajectories.

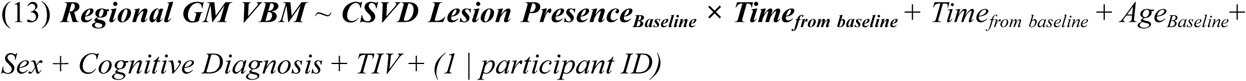

These models included participant-specific random intercepts and were adjusted for age, sex, cognitive diagnosis, and TIV. FDR correction for multiple comparisons was applied across all analyses, with a statistical significance threshold of *q_FDR_* < 0.05.

## 3 RESULTS

### 3.1 Participant characteristics

Participants were classified as stable CN (*n = 81*), stable SCD (*n = 83*), stable MCI (*n = 61*), converters (converter-SCD and converter-MCI combined; *n = 43*), or stable AD (*n = 21*; Table 1). Age and sex distribution differed across groups (*p < .001*). The stable CN and SCD groups were younger and included higher proportions of females than the stable MCI, converter, and stable AD groups. Educational attainment did not differ across groups (*p = .39*).

Most vascular health indicators did not differ significantly across groups. Neck, hip and waist circumferences were analyzed separately by sex. Among females, neck and waist circumference differed across groups (*p < 0.03*), whereas neither differed among males (*p > .05*). Overall group differences were also observed in HbA1c (*p = .04*), total and LDL cholesterol (*p = .002*), and HDL cholesterol (*p = .01*). The Hachinski Ischemic Score differed across groups (*p* < .001), with higher scores in the stable MCI group than in the stable CN and SCD groups and in the cognitive-decline group than in the stable CN group. The unweighted vascular composite score also differed across groups (*p = .004*), with higher scores in the stable MCI and converter groups than in the stable CN and SCD groups. The Framingham risk score did not differ significantly across groups (*p = .06*).

CSVD lesion burden was compared across the five diagnostic groups. Significant overall group differences were observed in PVS count (*p = .006*), PVS volume (*p = .04*), and WMH volume (*p < .001*). The stable MCI group had greater PVS count (*q_FDR_ = .009*) and volume (*q_FDR_ = 0.04*) than the stable CN group. WMH volume was greater in the stable MCI group than in the stable CN and SCD groups and greater in the stable AD group than in the stable CN, SCD, and converter groups. No other post hoc pairwise differences were significant. CMB count, presence, and volume did not significantly differ across groups.

### 3.2 Associations of vascular health indicators with CSVD lesions

In cross-sectional analyses, hypertension (*β = 0.24, p = .03*) and higher BMI (*β = 0.14, p = .01*), systolic blood pressure (*β = 0.12, p = .02*), diastolic blood pressure (*β = 0.13, p = .02*), HbA1c (*β = 0.12, p = .03*), and Framingham risk scores (*β = 0.33, p = .01*) were associated with greater PVS count. Higher HDL cholesterol was associated with lower PVS count (*β = -0.19, p<.001, q_FDR_=.02*) but greater WMH volume (*β = 0.11, p = .04*). Hypertension (*OR = 2.55, 95% CI: 1.52-4.26, p<.001, q_FDR_ = .008*), physical inactivity (*OR = 2.42, 95% CI: 1.27-4.63, p=.01*) and higher vascular composite score (*OR = 1.41, 95% CI: 1.10-1.81, p = .01*) were associated with greater odds of CMB presence. Higher total (*OR = 0.76, 95% CI: 0.58-0.99, p = .04*) and HDL (*OR = 0.76, 95% CI: 0.58-0.99, p =.04*) cholesterol were associated with lower odds of CMB presence (Figure 2A and eTable 1).

**Figure 2.**
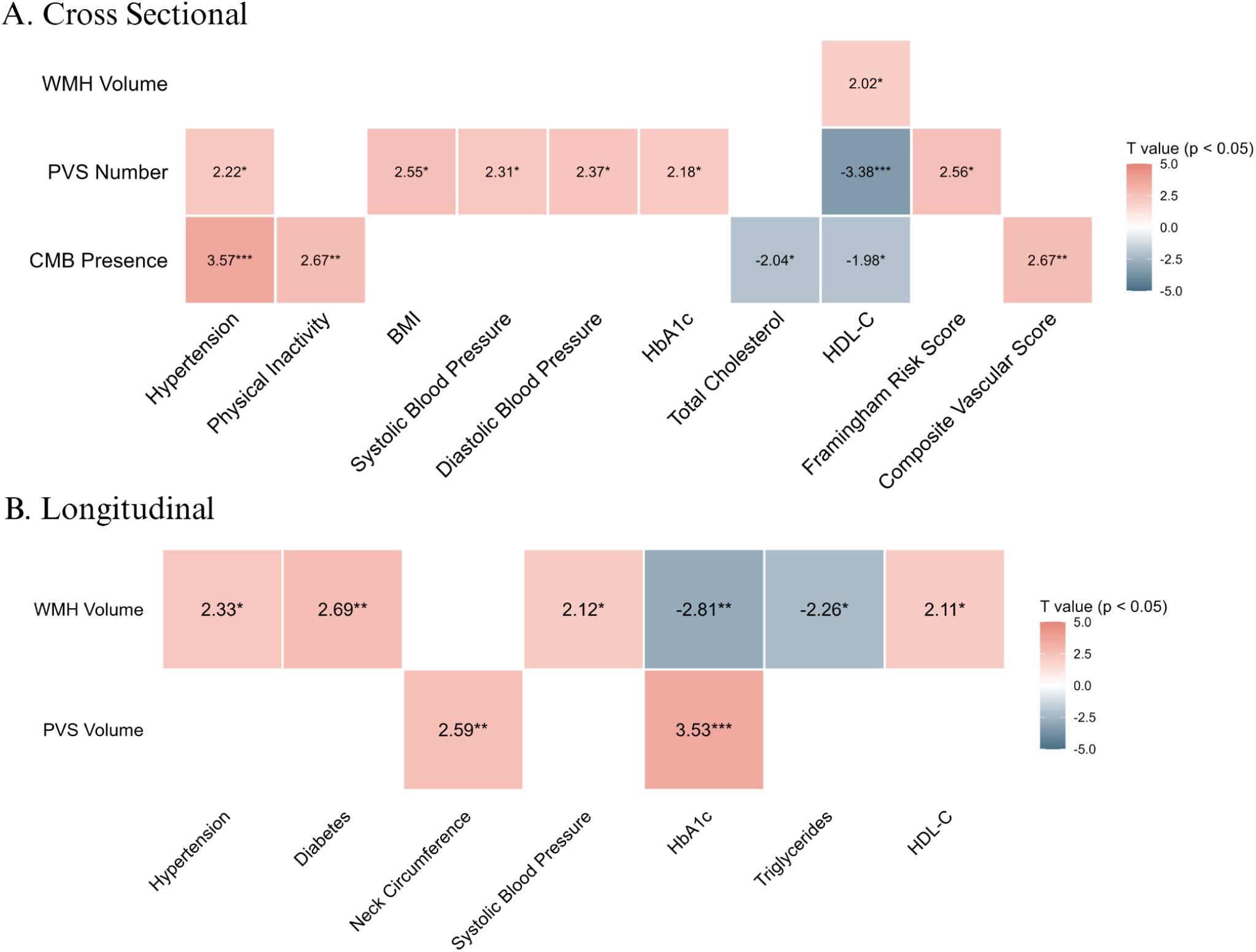
Significant cross sectional (panel A) and longitudinal (panel B) associations between vascular health indicators and CSVD measures. Associations not included did not reach the significance threshold of *p*<0.05. PVS, perivascular spaces; CMB, cerebral microbleeds; LAC, lacunae; BMI, body mass index; HbA1c, glycated hemoglobin; HDL-C, high-density lipoprotein cholesterol. TG: triglyceride. *: p<0.05; **:p<0.01, ***: p<0.001

In longitudinal analyses, hypertension (*β = 0.08, p = .02*) and diabetes (*β = 0.20, p = .01*) were associated with greater increases in WMH volume over time, as indicated by significant risk-factor-by-time interactions. Across repeated assessments, higher systolic blood pressure (*β = 0.05, p = .03*) and HDL cholesterol (*β = 0.09, p = .04*) were associated with greater WMH volume, whereas higher HbA1c (*β = -0.07, p<.001*) and triglycerides (*β= -0.08, p = .02*) were associated with lower WMH volume. Higher HbA1c (*β = 0.11, p < .001, q_FDR_=0.01*) and greater neck circumference (*β = 0.10, p = .01*) were associated with greater PVS volume (Figure 2B). No other associations reached statistical significance (eTable 2).

### 3.3 Associations among CSVD lesions

In cross-sectional analyses, WMH and PVS volumes were positively associated (*β = 0.33, p<.001*). Higher WMH volume (*OR = 1.79, 95% CI: 1.18-2.75, p=.007*) and higher PVS volume (*OR = 2.30, 95% CI: 1.49-3.69, p<.001*) were each associated with greater odds of lacune presence. Participants with lacunae had higher WMH volume (*β = 0.42, p=.01)* and higher PVS volume (*β = 0.61, p < .001*) than participants without lacunae. Lacune presence was also associated with greater odds of CMB presence (*OR = 2.10, 95% CI:1.00-4.49, p = .04*). No other cross-sectional (Figure 3, eTable 3) or longitudinal (eTable 4) associations reached statistical significance.

**Figure 3.**
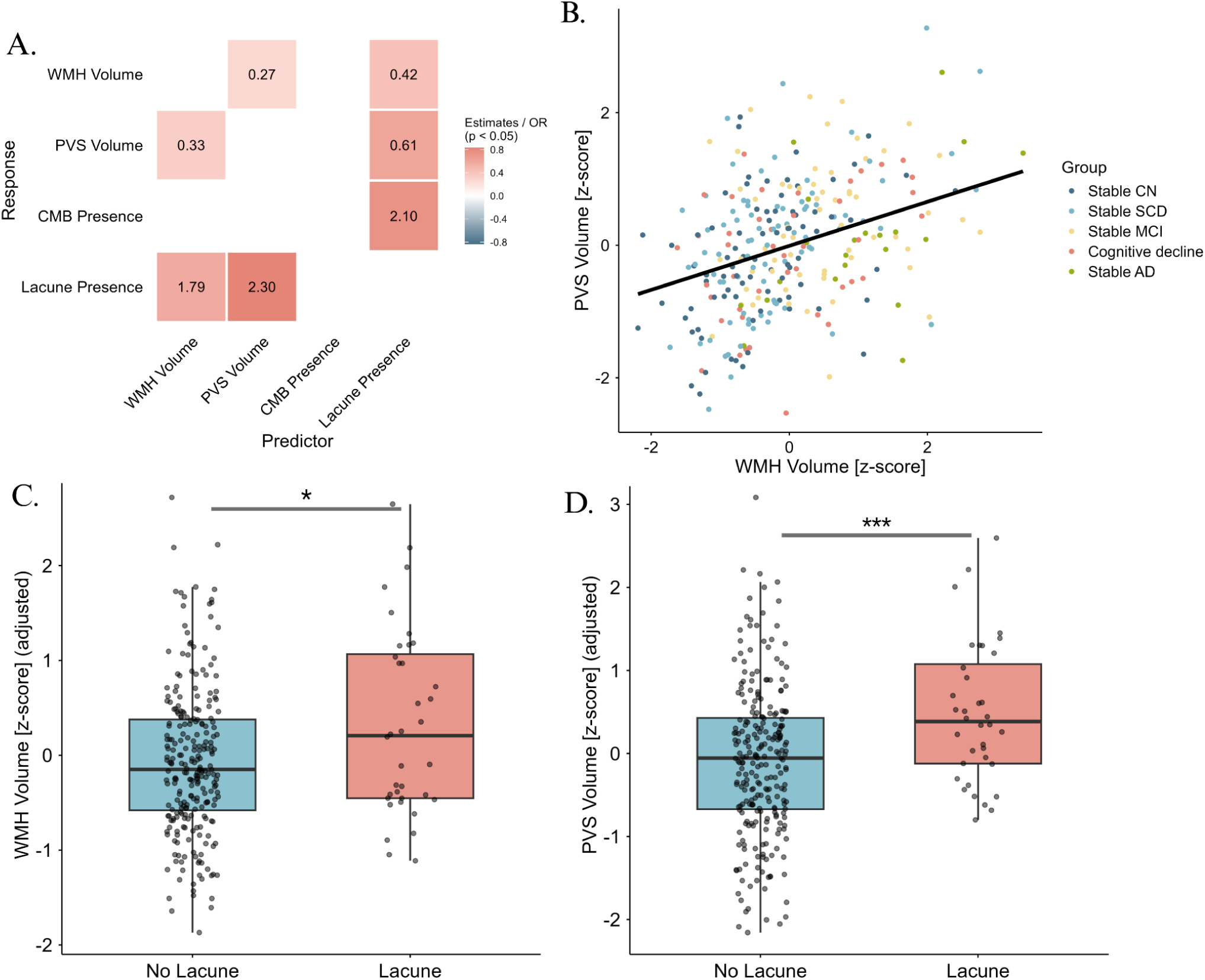
Cross-sectional associations among CSVD measures. (A) Effect estimates for significant associations among CSVD measures, presented as β coefficients for continuous outcomes and odds ratios (ORs) for binary outcomes. (B) Association between adjusted PVS and WMH volumes. (C) Difference in adjusted WMH volume between participants with and without lacunae. (D) Difference in adjusted PVS volume between participants with and without lacunae. *FDR-adjusted p < 0.05; **FDR-adjusted p < 0.01; ***FDR-adjusted p < 0.001.

### 3.4 Longitudinal trajectories of CSVD lesions

Longitudinal analysis included 153 participants who contributed 459 visits over a mean follow-up of 4.47 ± 2.37 years. Longitudinal trajectories were examined for WMH and PVS volumes (Figure 4). WMH volume increased significantly over time in the stable CN reference group (*β = 0.14, p<.001*). Compared with the Stable CN group, the converter group exhibited more rapid WMH accumulation (time-by-group interaction: *β = 0.20, p<.001*). Rates of WMH change did not differ significantly between the stable CN group and the stable SCD or stable MCI groups. PVS volume also increased significantly over time in the stable CN group (*β = 0.13, p = .006*). None of the time-by-group interactions were significant (all *p* > .05), indicating no evidence of differences in PVS trajectories across diagnostic groups (eTable 5).

**Figure 4.**
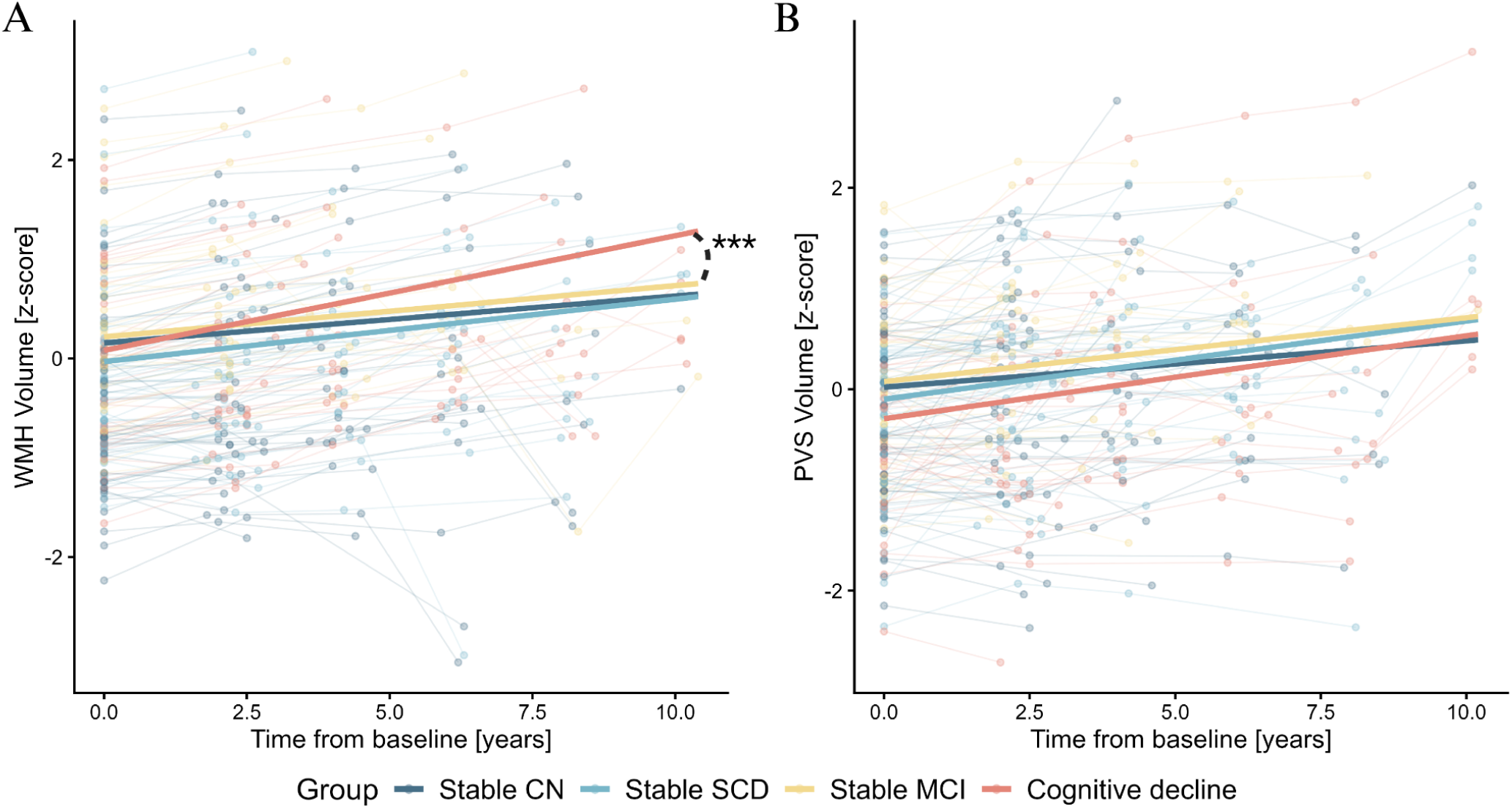
Longitudinal trajectories of WMH volume (Panel A) and PVS volume (Panel B). Points represent individual observations and are colored by the diagnostic group; repeated observations from the same participant are connected by lines. *: p<0.05; **:p<0.01, ***: p<0.001

### 3.5 Associations of CSVD lesions with cognition

At the first available assessment, performance on all cognitive measures differed across diagnostic groups (all *p<.001*, Table 1). The stable CN and SCD groups generally demonstrated the best performance across measures of global cognition (MoCA and T-MMSE), verbal learning and memory (RAVLT), language (BNT), processing speed and executive function (TMT part A and B and the TMT B/A ratios). The stable AD group generally demonstrated the poorest performance, while the stable MCI and converter groups showed intermediate performance.

In cross-sectional analyses, greater WMH volume was associated with lower MoCA scores (*β = -0.12, p = .01, q_FDR_ = .04*, Figure 5B) and poorer TMT-B performance (*β = -0.18, p =. 002, q_FDR_ = .02*, Figure 5B). Baseline lacune presence was associated with less favorable change in TMT-B performance (*β = -0.28, p = .002, q_FDR_ = .02*) and TMT B/A ratio (*β = -0.33, p = .004,q_FDR_ = .02*) over time (Figure 5D).

**Figure 5.**
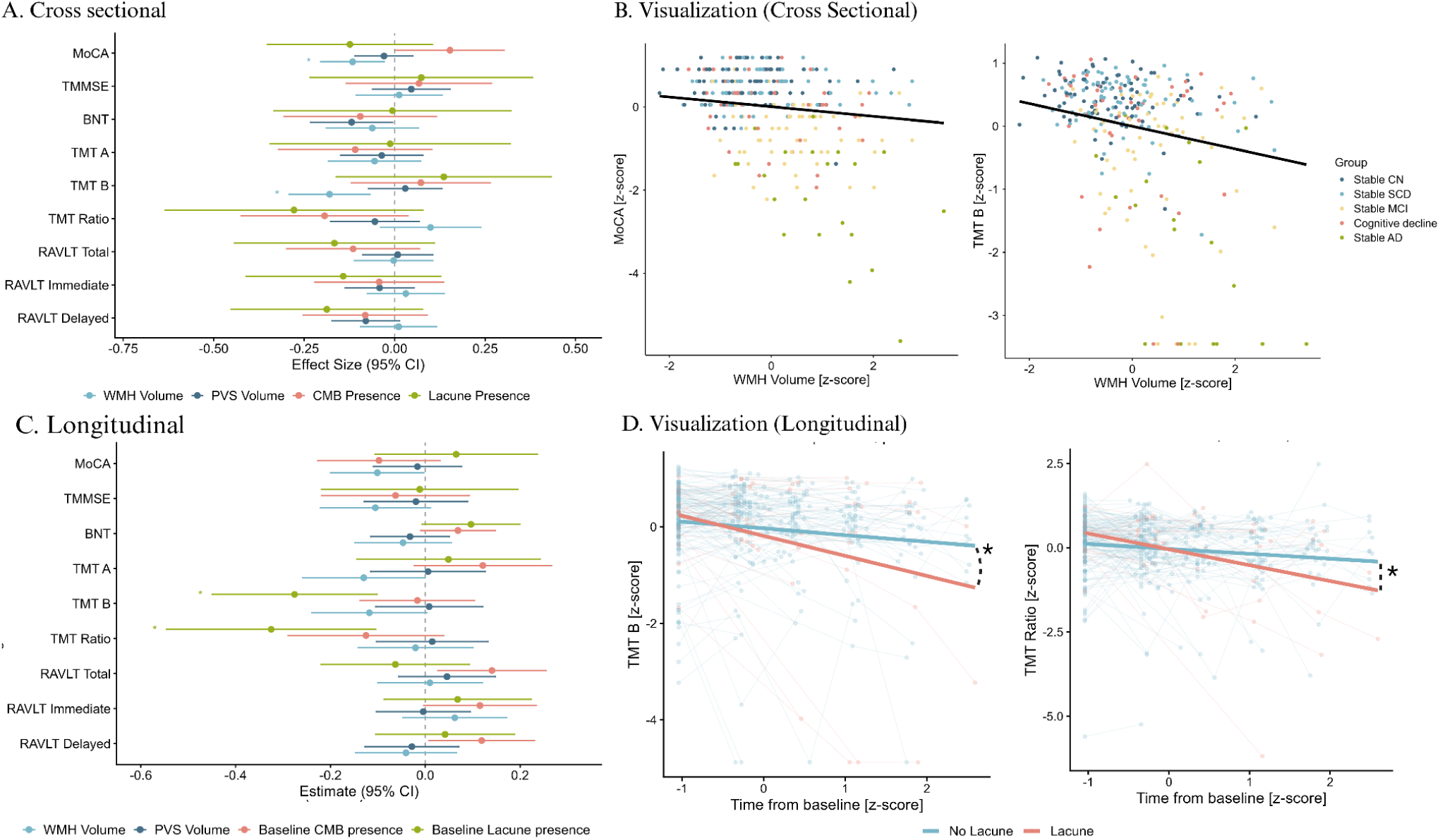
Associations between cognition and CSVD measures. β coefficients for cross-sectional (A) and longitudinal (C) associations between cognitive measures and CSVD measures. (B) Visualization of significant cross-sectional associations between MoCA and WMH volume, TMT-B and WMH volume (left to right). (D) Visualization of significant longitudinal associations between reversed TMT-B and lacune presence, reversed TMT-ratio and lacune presence (left to right). All TMT measures were reverse coded to match the direction of other cognitive tests. *FDR-adjusted p < 0.05; **FDR-adjusted p < 0.01; ***FDR-adjusted p < 0.001.

### 3.6 Associations of CSVD lesions with regional GM volumes

In cross-sectional analyses, greater WMH volume was associated with lower GM volumes in 21 regions (*β range = -0.24 to -0.16, q_FDR_<.05*), including the bilateral insula, basal forebrain, pars opercularis, cuneus, superior temporal, and supramarginal regions; the right lingual, postcentral, middle temporal, precuneus, and posterior cingulate regions; and the left rostral middle frontal, transverse temporal, pars triangularis, and superior frontal regions. In contrast, greater WMH volume was also associated with greater caudate volume bilaterally (*β range = 0.20 to 0.22, q_FDR_ < .02*). Greater PVS volume was associated with larger GM volumes in the right parahippocampal and caudate, and bilateral putamen regions (*β range = 0.19 to 0.24, q_FDR_<.02*; Figure 6A and eTable 8).

**Figure 6.**
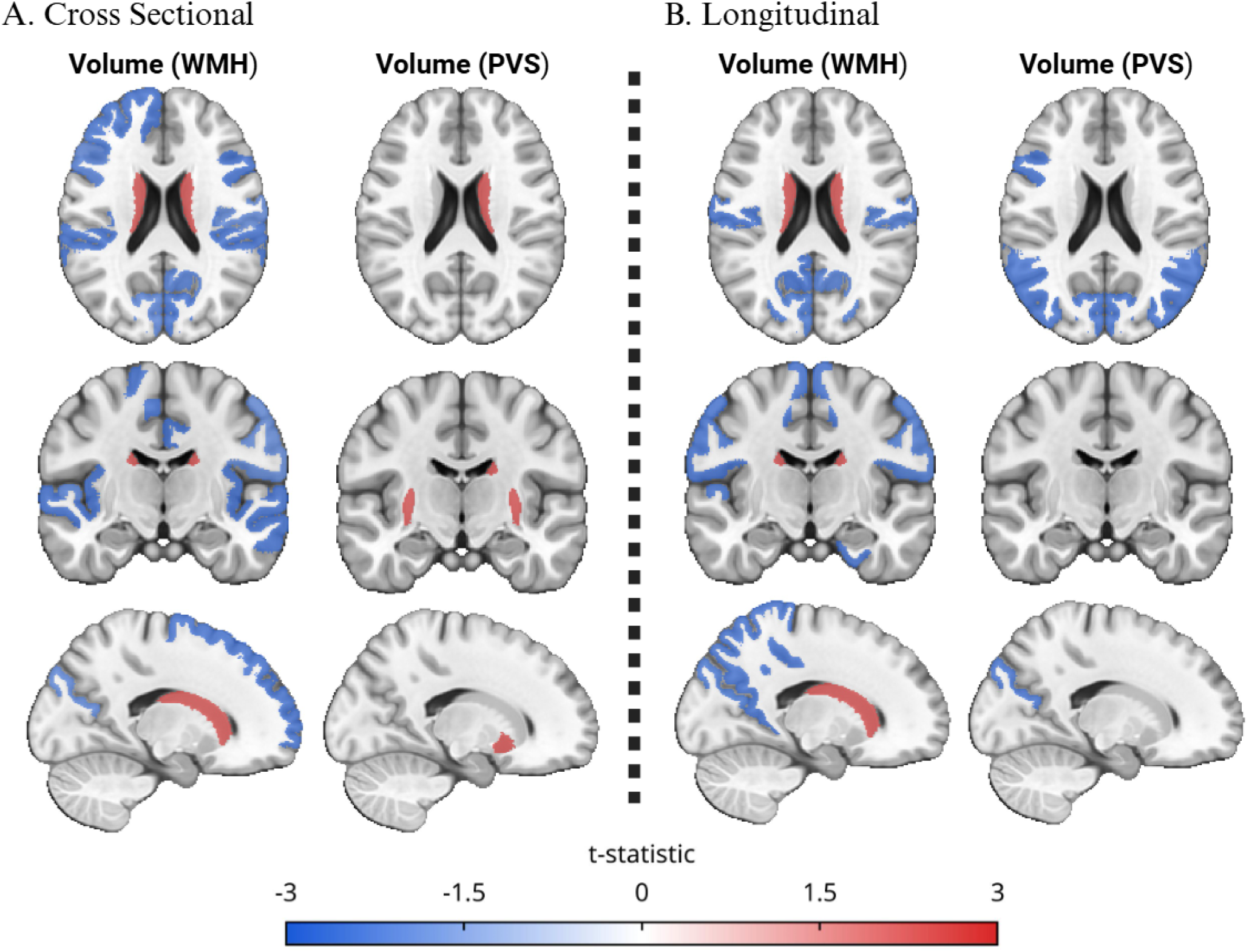
Associations between regional gray matter VBM measures and CSVD measures. T-values for cross-sectional (A) and longitudinal (B) associations between regional gray matter voxel-based morphometry (VBM) measures and CSVD measures. A shared color scale for T-values is shown at the bottom.

In longitudinal mixed-effects analyses, greater PVS volume was associated with lower GM volumes across repeated assessments in the bilateral inferior parietal and cuneus regions, bilateral cerebellar vermal lobules VI-VII, left pars opercularis, left pars orbitalis, and left cerebellar vermal lobules I-V (*β range= -0.06 to -0.05, q_FDR_<.05*). Greater WMH volumes were also associated with lower GM volumes in the bilateral superior parietal, postcentral, paracentral, and precuneus regions; right entorhinal region and cerebellar vermal lobules VIII-X; and left transverse temporal, isthmus cingulate, and cuneus regions (*β range= -0.09 to -0.05, q_FDR_ ≤.04*). Conversely, greater WMH volume was associated with greater caudate volume bilaterally (*β range= 0.10 to 0.12, q_FDR_ ≤ .002*).

## 4 DISCUSSION

The present study investigated the prevalence of and inter-relationships between CSVD lesions, as well as their associations with risk factors, neurodegeneration, and cognitive decline in a longitudinal cohort of aging adults on the spectrum of AD. To our knowledge, this is the first study examining the prevalence, burden, inter-relationships, and cognitive and neurodegenerative correlates of the four markers of CSVD in the same cohort. At baseline, WMH burden was greater in participants with MCI and AD, and PVS counts were higher in MCI, whereas CMB and lacunar infarct burden did not differ between diagnostic groups. Importantly, vascular risk factors including blood pressure, hypertension, BMI, cholesterol, triglycerides, and HbA1c, were associated not only with cross-sectional CSVD burden but also with greater lesion progression over time, especially for WMH burden and PVS volumes. These associations were lesion-specific, suggesting that distinct vascular risk profiles may contribute differentially to the development and progression of individual CSVD burden. Additionally, we found that different CSVD markers co-occurred with each other and WMH and PVS progression were associated with decreased GM volume over time. CSVD markers were also associated with both the presence and rate of cognitive decline across global and domain-specific measures. WMHs were associated with global cognition and executive function, while PVS were related to language impairment. Taken together, these findings support a role for CSVD pathology in both cognitive impairment and its progression within the AD spectrum, while highlighting substantial heterogeneity across individual CSVD markers, both in their association with cardiovascular burden and cognitive status and subsequent rates of cognitive decline.

The greater WMH burden in MCI and AD is consistent with previous work showing that WMH are common in individuals with cognitive impairment and are associated with both MCI and AD^23,60^, supporting the contribution of cerebrovascular injury to the clinical expression of AD^61^. The absence of group differences in CMBs and lacunae may reflect their previously observed lower prevalence^29,62^. Compared with estimates from older cohorts of broadly similar age, WMH volumes in the CIMA-Q CN and SCD groups were generally within the range of previously reported values, whereas greater volumes were observed in the MCI and AD participants^63,64^. Lacune prevalence was broadly consistent with previous reports from comparable older populations^65,66^, while PVS count and volume appeared higher than those reported in the Lothian Birth Cohort^63^. CMB prevalence in CIMA-Q was higher than the estimates reported in population-based aging cohorts^67–69^, although it was close to the higher prevalence reported in a cohort assessed using susceptibility-weighted imaging^70^.

The cross-sectional associations between vascular risk factors and CSVD burden further support the contribution of vascular health to the accumulation and progression of CSVD lesions. Blood pressure and hypertension, as well as metabolic factors including BMI, cholesterol, triglycerides, and HbA1c, were associated with CSVD burden and/or progression in our analyses. These findings are broadly consistent with longitudinal studies demonstrating associations between vascular risk and CSVD markers. The lack of association with composite risk scores such as the Framingham Risk Score may relate to how these scores were developed and the populations for which they were intended. The Framingham risk score was examined as a complementary measure as it provides clinically derived weighting of vascular risk factors but was originally developed for adults aged 30-74 years who were free of cardiovascular disease at baseline, criteria that were not met by all participants in this older cohort.

Our finding of broad associations among CSVD markers suggest that these imaging features represent overlapping, rather than independent, manifestations of small vessel pathology. Specifically, WMH, PVS, and lacunar infarcts tended to co-occur, whereas CMBs were more selectively associated with lacunar infarcts. This pattern may reflect common underlying small vessel pathology, while the differing associations among individual markers and with vascular risk factors may reflect distinct pathophysiological mechanisms or stages of vascular injury. For example, WMH and PVS may represent more diffuse manifestations of chronic small vessel dysfunction, whereas lacunar infarcts reflect more focal occlusive injury. The association between CMBs and lacunar infarcts may similarly reflect the coexistence of hemorrhagic and ischemic consequences of small vessel pathology. As few studies assessed all four markers of CSVD simultaneously and post-mortem evidence is lacking, these findings warrant further investigation.

The association of greater WMH and PVS burden with increased cortical atrophy provides evidence for a regional relationship between CSVD and neurodegeneration. This is notable as few studies have investigated this association and prior work has relied on visual ratings of vascular lesions and global atrophy^71–73^, which limit quantitative analyses and the characterization of their spatial relationships. In contrast, the positive associations of WMH burden with caudate volume and of PVS burden with putamen volume were unexpected. One potential explanation is that these findings reflect measurement errors rather than true increases in subcortical tissue volume. Previous work has shown that WMH can be systematically misclassified as GM in T1w MRI segmentation, particularly in periventricular regions such as the caudate, resulting in overestimation of caudate volumes^74^. Given their spatial overlap and that PVS share similar signal characteristics (i.e. low intensity on T1w MRI), a similar effect may have contributed to the positive association between PVS burden and putamen volume.

The observed associations between CSVD markers and both baseline cognitive impairment and longitudinal cognitive decline further support the clinical relevance of cerebrovascular injury in AD. Notably, these relationships were also lesion-specific, with WMHs showing cross-sectional associations with global cognition and executive function, and presence of lacunae at baseline being associated with longitudinal decline in executive function. These findings are consistent with previous evidence showing robust relationships between CSVD lesions and in particular WMH burden and executive function and global cognition^12,75,76^.

Our study has several strengths. Our longitudinal study design, including consistently acquired multi-modal MRI protocols, allowed for reliable characterization of risk factors, CSVD lesions, neurodegeneration, and cognition over a period of ten years. The neuroimaging pipelines that were used have been specifically designed and validated for use in multi-center and multi-modal datasets^46,49,51,77^. Furthermore, all image processing steps were followed by comprehensive quality control and CSVD lesion segmentations were corrected when necessary, ensuring robustness and accuracy of the derived metrics.

Some limitations warrant consideration. The relatively small sample size and limited follow-ups for the AD group limited our statistical power. CIMA-Q is a predominantly Quebec-based, racially and ethnically homogeneous, highly educated volunteer cohort that may have a lower vascular-risk burden than broader older populations. Recruitment through research centres may also underrepresent individuals facing barriers to healthcare, although medical care access and vascular-risk treatment were not directly assessed. Together, these characteristics may limit sample diversity and generalizability to other populations. Furthermore, our analyses were limited to global measures of CSVD burden and did not account for the spatial distribution of individual lesions. The anatomical location of CSVD markers may influence their clinical relevance, as well as their associations with specific vascular risk factors. Thus, incorporating the anatomical location of lesions in future studies may provide a more nuanced understanding of the relationship between vascular risk factors, CSVD progression, and cognitive outcomes.

In conclusion, our findings demonstrate that CSVD is common across the AD spectrum and that individual CSVD markers show distinct associations with vascular risk factors, neurodegeneration, and cognitive decline. The frequent co-occurrence of CSVD markers, together with the associations of WMH and PVS with cortical atrophy and cognitive impairment, supports a role for cerebrovascular pathology in neurodegeneration and cognitive decline. Overall, these findings highlight the heterogeneity of CSVD and the importance of considering all individual markers when investigating their clinical consequences.

## Data Availability

Data used in the preparation of this article were obtained from the Consortium for the early identification of Alzheimer's disease -Quebec (CIMA-Q; cima-q.ca). A list of researchers involved in the design of CIMAQ can be found on the cima-q.ca website. These researchers contributed to the establishment of protocols, the implementation of the research infrastructure, the recruitment and follow-up of participants, the obtaining of data, the maintenance of biological and ex-vivo samples, and certain derived data.

https://www.cima-q.ca/en/home/

## DATA AND CODE AVAILABILITY STATEMENT

CIMA-Q data can be accessed at http://www.cima-q.ca/en/home/. PELICAN is available at https://github.com/VANDAlab/Preprocessing_Pipeline. ShiVA tools are available at https://github.com/pboutinaud/SHiVAi/.

## Funding

ZC reports receiving funding from China Scholarship Council. AM receives doctoral scholarships from the Fonds de Recherche du Québec - Santé (FRQS, https://doi.org/10.69777/344731) and the Vascular Training (VAST) Platform. SR receives doctoral scholarships from the FRQS (https://doi.org/10.69777/2007562), VAST, and Brain Health (BH) Care. RM receives a doctoral scholarship from the FRQS (https://doi.org/10.69777/350961). KC receives a Canada Graduate Research Scholarship, Master’s, from the Canadian Institutes of Health Research (CIHR) and a doctoral scholarship from VAST. YZ reports receiving research funding from the FRQS (https://doi.org/10.69777/379799), Natural Sciences and Engineering Research (NSERC), and CIHR. MD reports receiving research funding from the CIHR (191303, 198104, and 213169), NSERC discovery grant (RGPIN-2023-04038), FRQS (https://doi.org/10.69777/330750), Alzheimer Society Research Program (ASRP), and Tier-2 Canada Research Chair in Vascular and Neurodegenerative Disorders of Aging.

## Declaration of Competing Interests

The authors report no competing interests.

## ACKNOWLEDGEMENTS

The authors acknowledge Digital Research Alliance of Canada (https://www.alliancecan.ca/en) for the usage of the computing resources in the current work. The CIMA-Q investigators contributed to the design, protocols and implementation of the study, as well as the collection of clinical, cognitive and neuroimaging data and biological samples. A complete list of the CIMA-Q investigators can be found at www.cima-q.ca. The CIMA-Q is supported by the Fonds de recherche du Québec-Santé (FRQS) - Pfizer Innovation Program, FRQ cohort funds, the Quebec Network for Research on aging, the Fondation Courtois (NeuroMod project), the Consortium for the Neurodegeneration associated with Aging, CIHR and the Fondation Famille Lemaire.

## Notes

### Competing Interest Statement

The authors have declared no competing interest.

### Author Declarations

The Research Ethics Board (REB) of the Centre integre universitaire de sante et de services sociaux de l'Ouest-de-l'Ile-de-Montreal - Mental Health and Neuroscience Subcommittee gave ethical approval for this work.

